# The impact of the COVID-19 pandemic on children with medical complexity

**DOI:** 10.1101/2021.12.02.21266765

**Authors:** Catherine Diskin, Francine Buchanan, Eyal Cohen, Tammie Dewan, Tessa Diaczun, Michelle Gordon, Esther Lee, Charlotte Moore-Hepburn, Nathalie Major, Julia Orkin, Hema Patel, Peter J. Gill

**Author notes:** **Corresponding Author**: Dr. Catherine Diskin, Black Wing 8227, Division of Paediatric Medicine, The Hospital for Sick Children, 555 University Avenue, Toronto, Ontario, M5G 1X8, Contact Number. **Ethics statement:** Consent (from patients) is not required for public health surveillance purposes. We are able to collect the information without consent on behalf of PHAC under the Department of Health Act and the Public Health of Agency Act. **Consent statement:** Physicians (paediatricians and paediatric subspecialists) provide their consent when participating in the Canadian Paediatric Surveillance Program. **Contributor’s statement**. Catherine Diskin and Peter Gill conceptualized and developed the study, convened the steering committee, carried out initial analyses, drafted and revised the initial manuscript. Dr. Peter Gill guided the study design. Francine Buchanan, Eyal Cohen, Tammie Dewan, Tessa Diaczun, Michelle Gordon, Esther Lee, Natalie Major, Charlotte Moore-Hepburn, Julia Orkin and Hema Patel were members of the study steering committee. All authors contributed to developing the study protocol, questionnaire designed and interpretation of results. All authors approved the final manuscript as submitted and agree to be accountable for all aspects of the work.

## Abstract

**Background:** Descriptions of the COVID-19 pandemic’s indirect consequences on children are emerging. We aimed to describe the impacts of the pandemic on children with medical complexity (CMC) and their families.

**Methods:** A one-time survey of Canadian paediatricians using the Canadian Paediatric Surveillance Program (CPSP) was conducted in Spring 2021.

**Results:** A total of 784 paediatricians responded to the survey, with 70% (n=540) providing care to CMC. Sixty-seven (12.4%) reported an adverse health outcome due to a COVID-19 pandemic-related disruption in healthcare delivery. Disruption of the supply of medication and equipment was reported by 11.9% of respondents (n=64). Respondents reported an interruption in family caregiving (47.5%, n=252) and homecare delivery (40.8%, n=218). Almost 47% of respondents (n=253) observed a benefit to CMC due to COVID-19 related changes in healthcare delivery, including increased availability of virtual care and reduction in respiratory illness. Some (14.4%) reported that CMC were excluded from in-person learning when their peers without medical complexity were not.

**Conclusion:** Canadian paediatricians reported that CMC experienced adverse health outcomes during the COVID-19 pandemic, including disruptions to family caregiving and community supports. These results highlight the need for healthcare, community and education policymakers to collaborate with families to optimize their health.

**“What This Study Adds”:**

- Children with medical complexity experienced adverse health outcomes related to the direct and indirect effects of the COVID-19 pandemic.
- The COVID-19 pandemic has interrupted family caregiving, homecare support, access to education, and key supports for CMC and their families.
- Canadian paediatricians observed benefits associated with structural changes relating to the COVID-19 pandemic, including the expansion of virtual care and the reduced incidence of respiratory illness

## INTRODUCTION

Children, including those with chronic illness, have been less affected by severe acute respiratory syndrome coronavirus 2 (SARS-CoV-2) infection than adults, with a small proportion of documented coronavirus disease 19 (COVID-19) cases, hospitalizations and deaths affecting children.^1–3^ The pandemic has resulted in rapid change across many aspects of society, including healthcare and education.^1^ Descriptions of the pandemic’s indirect consequences on children are emerging, including the impact on children with special health care needs, particularly those with medical complexity.^4^ Children with medical complexity (CMC) have special healthcare needs with chronic conditions resulting in caregiver burden, technology dependence, and frequent healthcare interactions across multiple settings, including tertiary and community hospitals, primary care settings, schools, and communities.^5^ Family caregivers perform complex medical and therapy-related tasks supporting the delivery of care.^6,7^

As a direct consequence of COVID-19 pandemic restrictions, CMC have experienced multiple disruptions in their care,^4^ including disruption of medical and specialist care, therapy and rehabilitative services, homecare and respite services and education.^8^ For example, disruption to school can interrupt the provision of developmental services, out-of-home respite and increase the caregiving demands of parents and caregivers.^1^ Further disruption in income, financial supports, transportation and the supply of vital equipment and supplies (e.g. medicines, specialized formulas) provided additional challenges. However, no prior study has examined these effects and adverse outcomes associated with the disruption of services to CMC during the COVID-19 pandemic.

We aimed to describe the indirect impacts of the COVID-19 pandemic on CMC and their families, focusing on (1) medical services, including access to and family experience, (2) community services including home care, respite, and timely access to medication and medical equipment and (3) education, including access to in-person or virtual learning environments, and disruptions to in-school delivery of nursing and therapies.

## METHODS

We conducted a one-time survey of Canadian paediatricians and subspecialists using the Canadian Paediatric Surveillance Program (CPSP). The CPSP is a joint initiative of the Public Health Agency of Canada (PHAC) and the Canadian Paediatric Society (CPS). The program is responsible for conducting national surveillance into uncommon child and youth health conditions low in frequency but high in associated morbidity and mortality. The program surveys over 2800 paediatricians.

### Survey development

The survey was developed collaboratively by the CPSP Scientific Steering Committee, which included clinicians (physicians, nurse practitioners) providing care to CMC across various clinical settings, including academic, community and rural, and the parent of a child with medical complexity.

We asked respondents whether they provided care to CMC as part of their clinical practice; respondents who did completed the survey. The survey collected information on respondent demographics, healthcare delivery, including experiences of adverse health events related to a COVID-19 pandemic-related disruption in routine healthcare delivery, changes in healthcare that benefit CMC, the impact of COVID-19 pandemic on family caregiving, homecare, and supply of medication and equipment. Information about the educational experience of CMC during the pandemic, including virtual learning, school supports, in-school delivery of nursing and therapies, was collected. When respondents reported an indirect impact of the COVID-19 pandemic (e.g., disrupted homecare), they were asked the frequency of outcome or the percentage of CMC and families affected and the frequency of contributing factors. Appendix 1 contains the final copy of the 17-item survey.

### Survey distribution

The survey was distributed in English and French in February 2021, with a closing date in April 2021. Two reminders were sent to online respondents who had yet to responded.

### Analysis

Microsoft Excel (version 2015) was used to tabulate responses, and statistical analyses were descriptive. The study was funded through a CPSP in-kind grant. Data elements where the “n” value is less than 5 were reported according to CPSP policy.

## RESULTS

### Participants

A total of 784 paediatricians responded, a response rate of 27.7% (784/2826), including 770 online responses and 14 hard copy surveys. Of those who responded, 70% (n=540) provided care to CMC, and their responses are described below. Respondents represented all provinces and territories except Nunavut (Table 1). Most respondents were general paediatricians (58%; n=312) followed by subspecialists (40%; n=218).

**Table 1:** Characteristics of physician respondents who provide care to CMC (n=540)

| **Characteristic** | **% (n)** |
| --- | --- |
| *Type of physician\** |  |
| General paediatrician | 57.8% (312) |
| Subspecialist | 40.4% (218) |
| Missing | 1.9% (10) |
| *Location – provinces* |  |
| Atlantic Canada ** | 7.7% (42) |
| The Prairies** | 20.9% (113) |
| *Location - provinces* |  |
| Alberta | 13.3% (72) |
| British Columbia | 11.7% (63) |
| Manitoba | 5.2% (28) |
| New Brunswick | 1.9% (10) |
| Newfoundland and Labrador | 1.3% (7) |
| Northwest Territories | <5 |
| Nova Scotia | 4.4% (24) |
| Nunavut | 0 |
| Ontario | 36.9% (199) |
| Prince Edward Island | <5 |
| Quebec | 20.9% (113) |
| Saskatchewan | 2.4% (13) |
| Yukon | <5 |
| Missing | 1.5% (8) |
| *Practice location* |  |
| Urban | 71.7% (377) |
| Suburban | 12.1% (64) |
| Rural/remote | 8.9% (47) |
| Combined urban and suburban | 3.4% (18) |
| Combined urban and rural/remote | 1.8% (10) |
| Combined urban, suburban and rural/remote | 1.5% (8) |
| Combined suburban and rural/remote | <5 |
| Missing | 2.4% (13) |
| *Practice type* |  |
| Academic | 63.7% (344) |
| Non-academic | 36.2% (196) |
| Missing | 7.4% (41) |
| *Practice setting* |  |
| Combination - inpatient and outpatient | 18.7% (95) |
| Community – private office | 17.5% (89) |
| Inpatient | 13.6% (69) |
| Outpatient | 9% (46) |
| Combined inpatient, emergency department or urgent care centre and community – | 8.5% (43) |
| private office |  |
| Combined inpatient, outpatient and community – private office | 6.5% (33) |
| Combined inpatient, emergency department or urgent care centre and outpatient | 6.3% (32) |
| Emergency Department | 5.7% (29) |
| Combined inpatient and private office | 5.7% (29) |
| Other combinations | 8.5% (46) |
| Missing | 5.7% (29) |

\* Counts and percentages may be larger than the number of respondents due to multiple responses per respondent
\*\* Counts and percentages may be less than the number of respondents as we not report values <5

### Healthcare experience

#### Adverse health outcomes

One in nine respondents (12.4%; n=67) reported a CMC having an adverse health outcome due to a COVID-19 pandemic-related disruption in healthcare delivery, with responses from all provinces and regions (Table 2). Respondents reported 546 events of children who experienced an adverse outcome, with a median number of events of 3 (interquartile range [IQR] 1-10) per respondent. The most-reported observed adverse health outcomes are outlined in Table 2. Respondents highlighted delayed presentation to healthcare or deferral of care because of the risk of exposure to SARS-CoV-2 at a tertiary care centre, observed slowing of developmental gains, reduced provision of health surveillance, delays in accessing elective procedures and challenges associated with hospital visitation policies. There were no reported deaths.

**Table 2:** Experiences of CMC and their families during the COVID-19 pandemic as reported by Canadian paediatricians.

| **Adverse health outcomes experienced by CMC** |  |
| --- | --- |
| *Adverse health outcomes* | *Number of respondents* |
| Hospital admissions | 26 |
| Loss of physical or developmental gains | 22 |
| Extended hospital admission | 12 |
| Intensive Care Unit admission | 6 |
| Unplanned surgery | <5 |
| Death | 0 |
| **Reasons provided by paediatricians as contributing to a particular experience** |  |
| *Interrupted family caregiving* | *Number of respondents* |
| School / respite service closure | 116 |
| Family caregiver mental illness or burnout | 70 |
| Interrupted rehabilitation | 49 |
| Loss of homecare | 45 |
| Increased financial stress | 32 |
| Family caregiver self-isolation (COVID-19 exposure or illness) | 24 |
| Other | 46 |
| *Interrupted homecare* |  |
| Decreased homecare availability | 135 |
| Family choice to discontinue or limit homecare | 60 |
| Homecare worker illness or need for self-isolation | 22 |
| Family self- isolation (COVID-19 exposure/ symptoms /infection) | 18 |
| Other | 7 |

#### Interruption of medication and equipment supply

Almost 12% (11.9%; n=64) reported a disruption in medication and other supplies during the COVID-19 pandemic, with 37% (n=199) reported no disruption, and the remainder (51.3%; n=277) were uncertain (Table 3). Respondents reported that medication, medical equipment and routine vaccination were most frequently impacted. The most frequently cited reasons included medication or supply shortage and delays in dispensing. The impact of difficulties accessing medication or equipment was minor (i.e., short delay, no change in care; n=27) or moderate (i.e., alternative medication sourced without change in clinical status; n=26). Seven respondents reported a significant event (i.e., an adverse clinical event), with five reporting that the event resulted in hospitalization. The complications due to the interrupted medication and equipment supply included ill-fitting equipment that required adjustment, including skin breakdown and pain. Other factors impacting medication and equipment supply included the loss of employment of family caregivers, which affected their ability to pay for medication and supplies, challenges with equipment that remained in school, and dispensing practice changed as clinics changed from in-person to virtual.

**Table 3:** Respondents reporting of the experience of CMC during the COVID-19 pandemic.

|  |  |  |  |
| --- | --- | --- | --- |
| *Healthcare system* |  |  |  |
|  | *Yes* | *No* | *Unknown* |
| Adverse health outcome | 67 (12.4%) | 87.6% (473) |  |
| Observed benefit of pandemic | 46.9% (253) | 48.3% (261) | 4.8% (26) |
| Interrupted family caregiving | 47.5% (252) | 27.0% (143) | 25.5% (135) |
| Disrupted home care | 40.8% (218) | 30.1% (161) | 29.1% (156) |
| Disrupted supplies | 11.9% (64) | 36.9% (199) | 51.0% (277) |
| *Education system* |  |  |  |
| Excluded from in-person learning | 14.4% (78) | 43.0% (232) | 42.6% (230) |
| Receipt of healthcare via education system | 66.1% (357) | 11.5% (62) | 22.4% (121) |
| Transfer of resources | 8.3% (45) | 31.3% (169) | 60.4% (326) |
| Excluded because of PHA advice | 8.3% (45) | 31.7% (171) | 60.0% (324) |

**Table 4:** Positive impacts observed by paediatricians during the COVID-19 pandemic as related to CMC and their families.

| **Table 4: Positive impacts observed by paediatricians during the COVID-19 pandemic as related to CMC and their families** |
| --- |
| **Virtual care** |
| *Benefits to family*<br>• Reduced travel time<br>• Reduced need for preparation, including arrangement of specialized equipment and transport<br>• Reduced travel costs<br>• Reduced caregiver time away from employment<br>• Reduced waiting room exposure (helpful for children with behavioural and sensory challenges)<br>• Multiple providers present, including those working at different locations |
| *Benefits to clinician*<br>• Assessment – observation of child’s developmental skills in home environment, opportunity to speak with family caregivers alone<br>• Team functioning – ease of virtual meetings |
| *System development*<br>• Technology to support receipt of videos<br>• Development of billing codes to support physicians providing virtual care, including telephone follow-up financially |
| **Observed reduction in encounters of CMC with a respiratory illness** |
| **Healthcare system change** |
| *Changes in patient flow*<br>• New clinical programs to meet demand (e.g. eating disorders)<br>• New clinical pathways (e.g. direct admission to a ward) |

#### Family caregiving and homecare delivery

Two-hundred and fifty-two respondents (47.5%) reported that they observed an interruption in family caregiving to CMC during the pandemic, while over 40% of respondents (n=218) reported that families with CMC experienced disrupted homecare. Of the respondents who reported disruptions to family caregiving and homecare delivery, Table 2 outlines the most frequent reasons.

#### Benefits in the healthcare experience of CMC as observed by paediatricians

Almost half of the respondents (46.9%; n=253) reported observing a benefit of COVID-19 related changes in healthcare experienced by CMC. Respondents highlighted the increased availability of virtual care (n=184), the observed reduction in incidence of respiratory illness among CMC (n=60) and healthcare system change (n=12). Table 3 outlines the benefits in the healthcare experience of CMC and their families as observed by paediatricians. When asked about virtual care, respondents highlighted that a virtual visit could not replace all in-person visits – emphasizing the importance of the clinical examination in monitoring disease progression (e.g., scoliosis).

The reasons provided for the observed reduction in CMC experiencing respiratory illness included improved universal infection control precautions, including in healthcare (e.g., reduced numbers in waiting rooms) and their introduction into education settings (e.g., masking), in addition to their siblings not experiencing intercurrent illnesses. Respondents noted that the absence of respiratory illness had promoted CMC’s developmental progress.

### Educational experience

Of the respondents who could provide details of CMC attending school in 2020 (n=376), one-quarter (23.6%; n=89) reported that >80% of CMC attended in-person school while over one-third (37%; n=114) reported that <20% did. Before 2020, 399 paediatricians estimated in-person school attendance, with 75% of respondents estimating that >80% of CMC in their practice attended in-person school before the pandemic, with few paediatricians (5.5%; n=22) reporting that less than 20% of CMC attended in-person school. The change in school attendance as observed by participating paediatricians is outlined in Figure 1.

**Figure 1:**
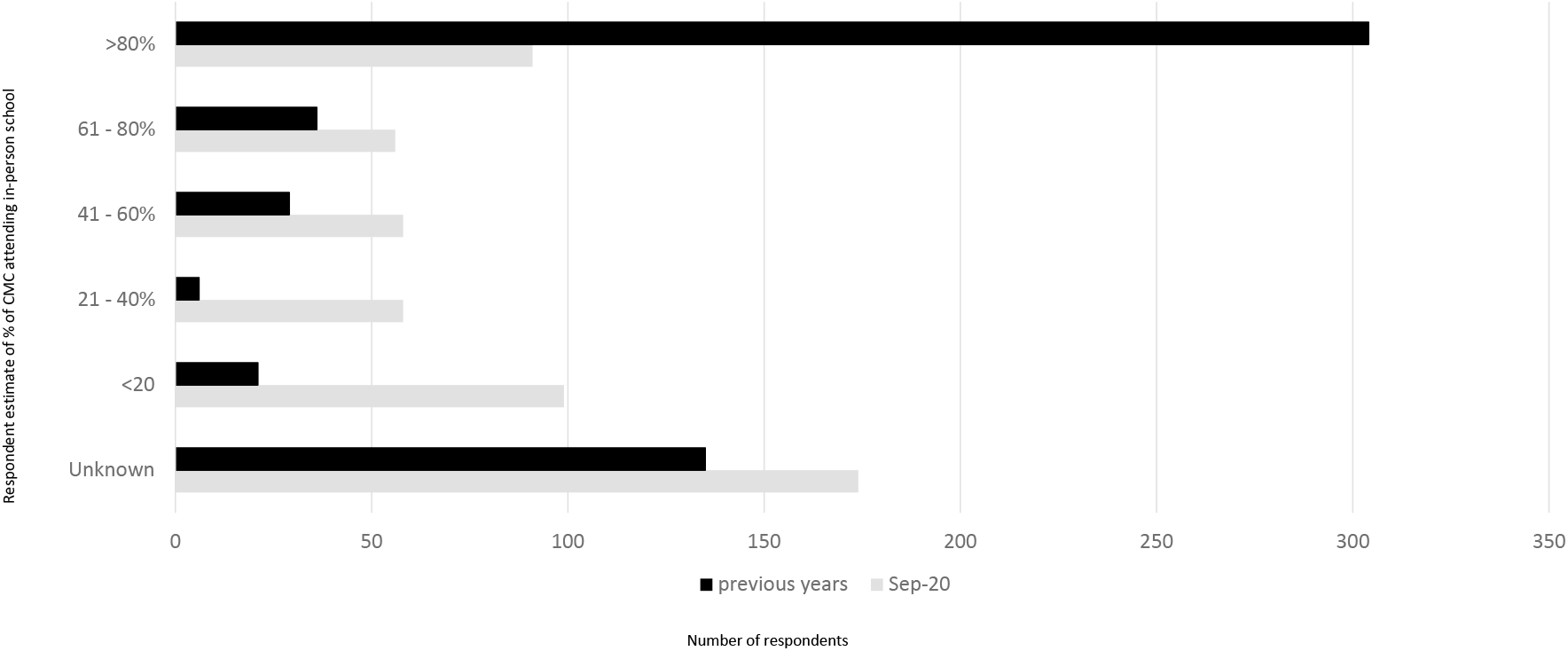
Percentage of school-aged CMC in your practice attended school in-person in September 2020 versus previous years

Seventy-eight respondents (14.4%) reported that CMC were excluded from in-person learning when their peers without medical complexity were not. Two hundred and thirty-two respondents (43%) reported no such experience, and 225 (41.7%) did not know. We invited respondents to share why CMC were excluded from in-person learning. Examples included public health advice that precluded their attendance, including advice regarding aerosol-generating medical procedures (AGMPs) and CMC being resident in a long-term care (LTC) facility, the absence of in-school behaviour support, challenges with transportation, physician recommendation and individual limitations in terms of maintaining physical distancing or mask wearing.

Two-thirds of respondents (66.1%; n=357) reported that CMC receive healthcare services at school. Very few respondents reported that services transferred from school to home and or community during periods of virtual learning (Table 3). Respondents described the loss of access to therapy and equipment that typically remains in school as having a deleterious impact on CMC. Respondents also highlighted that many families opted not to send CMC to school during the past year. Others highlighted that learning was impacted by the absence of and challenge of delivering an individualized education plan (IEP) during virtual school.

#### Regional differences in the indirect impacts of the COVID-19 pandemic

Regional differences in the indirect impacts of the COVID-19 pandemic are outlined in Appendix 2, including the percentages of respondents from each province. There were minimal discernable differences across provinces in responses.

## DISCUSSION

Canadian children with medical complexity experienced several indirect impacts of the COVID-19 pandemic. Our national survey identified adverse health outcomes and disruptions to family caregiving, available supports and school attendance.

Adverse health impacts resulting in CMC requiring both hospital admissions, including the intensive care unit (ICU) and an extended time in hospital, loss of physical or developmental and unplanned surgery mirrors previously published literature, highlighting disruption caused by the COVID-19 pandemic to safe, timely and effective care.^9^ CMC are a vulnerable population and it is important to recognise that they are disproportionately vulnerable to the consequences of the COVID-19 pandemic due to greater healthcare needs, dependency on community-based services and mental health concerns.^10^ Respondents highlight this reality for CMC and their families, with over 40% observing a disruption in community services, including homecare services. Homecare is essential to the lives of many CMC and their families,^11,12^ but limitations regarding its delivery were described before the pandemic.^13^ We also describe disruptions in family caregiving, highlighting the emotional and financial stress involved in caregivers of CMC.^14^ The impacts of the pandemic as related to family caregiving and community services interact with previously existing challenges, highlighting the vulnerability of this subgroup of children and their families and a pressing need to recognise the stressors CMC and their families are experiencing.

Canadian pediatricians reported major disruption to the educational system during the COVID-19 pandemic, including the well-known reduction in attendance at in-person class.^15^ School closures exert a greater influence on vulnerable children, including those with disabilities.^16^ Before the pandemic, many families of children with developmental disabilities, would highlight school holidays as times of increased stress.^17^ School is not simply an academic pursuit for many children, including CMC, but a place of therapy, nursing, respite, learning, and socialization. When access to such services are interrupted, it becomes the responsibility of family caregivers to step in. For families of CMC, this can be an additional burden to already overburdened caregivers. As the 2021-2022 academic year starts, considering the entire experience with an inclusive lens is necessary with collaboration and flexibility between traditional silos so that CMC can receive holistic care.

Almost half of the respondents highlighted some positive changes observed during the pandemic, including the expansion of virtual care and a reduction in children presenting with respiratory illnesses. The accelerated expansion of virtual care during the COVID-19 pandemic is previously described.^18^ The experience of respondents who reported a decrease in numbers of CMC presenting with respiratory illness supports empiric evidence.^19^Measures to mitigate the spread of SARS-CoV-2 likely contributed to a decline in other respiratory illnesses, including respiratory syncytial virus (RSV) and influenza.^20^ More recently, delayed seasonal surges in RSV have been described in both hemispheres, potentially correlating with the relaxation of some measures.^21,22^ The trend in reduced incidence of respiratory infections is particularly important to CMC, considering they experience greater rates of hospitalization and morbidities associated with such infections.^23–25^ The pandemic highlighted potentially valuable tools to protect the physical health of CMC. Formal evaluation of many interventions, including masking practice, hand washing, smaller class sizes, screening for respiratory symptoms, may have impacted transmission of respiratory illness, would provide an improved understanding of which measures lead to improved health outcomes and support safe and accessible service delivery for CMC and their families.

The observations of participating paediatricians on the experience of CMC and their families are far-reaching, crossing many aspects of CMC care and life. Their often-isolated experience with a continued need for advocacy was highlighted. For example, the impact of reduced support, such as family caregiving and home care, on families could exacerbate social isolation and loneliness they experienced before the pandemic, for which they receive little support.^26^ This study illustrates how the COVID-19 pandemic exacerbated an existing situation for CMC families and providers, who often must navigate silos.^26^ Over the past decade, there have been concerted efforts to coordinate CMC’s care across these sector silos.^27^ A pertinent example is how many CMC receive healthcare, including nursing and various therapies through the education system, and much of the related equipment remains in school. When in-person learning was interrupted, so did their access to healthcare, potentially interrupting their developmental progress. Few respondents reported that school-based resources were transferred elsewhere when schools remained closed. Another important factor that impacted school attendance for some CMC were challenges accessing transport. Many CMC require transportation to and from school, and when unavailable, often cannot attend school.

Our study has several limitations. First, information collected is based on voluntary reporting and is limited by a low response rate (27.7%), incomplete responses and recall bias. The response rate is in line with previous CPSP surveys. Second, respondents provided estimates of the numbers of families and children impacted by various aspects of the pandemic, limiting specific information. As such, we were unable to calculate population-based estimates. Third, we cannot verify that each response is unique, as there may be duplicate entries (e.g. physicians working in the same institution may describe the same event). Fourth, our survey was limited to paediatricians, and CMC receive care from many clinicians, including family physicians, nurse practitioners and therapists who could provide valuable insights. Fifth, paediatricians may often be unaware of all aspects of a child’s life, as evidenced by the high number of respondents that did not know the answer to particular questions. Finally, the survey did not include the experience of family caregivers, who are best placed to describe their children’s experience during the pandemic. Some indirect impacts on CMC may not have been visible to paediatricians due to the efforts of family caregivers, e.g., stockpiling medications and supplies in case medication or equipment shortages arose.

## CONCLUSION

Consideration of the broad impact on this sub-group of the paediatric population is required to inform pandemic and non-pandemic health policy and planning, including service design and delivery across acute, home and community care sectors, and education policy and service planning for children with CMC.

## Data Availability

All data produced in the present study are available upon reasonable request to the authors

## Abbreviations

AGMPs’: aerosol generating medical procedures
BC: British Columbia
COVID-19: coronavirus-19
CMC: child with medical complexity
CPSP: Canadian Paediatric Surveillance Program
IQR: interquartile range
LTC: long-term care
PPE: personal protective equipment
RSV: respiratory syncytial virus
SARS-CoV-2: severe acute respiratory syndrome coronavirus

## ACKNOWLEDGEMENTS

The study team acknowledges the time and contribution of the clinicians who participated in the study and Melanie Laffin Thibodeau who facilitated development and distribution of the survey.

