## Supplemental file 2 for "The impact of the COVID-19 pandemic on children with medical complexity"

**Appendix 2**: Respondents reporting of the experience of CMC during the COVID-19 pandemic (by province)

| Location | Canada*  (n=540) | Atlantic Canada** (n=42) | BC  (n=63) | Ontario  (n=199) | Quebec  (n=113) | Prairies***  (n=113) |
| --- | --- | --- | --- | --- | --- | --- |
| **Healthcare System** | | | | | | |
| Adverse health outcome | 12.4% (67) | 12% (5) | 16% (10) | 14% (28) | 8% (10) | 12% (14) |
| Observed benefit of pandemic | Y: 46.9% (253)  N: 48.3% (261)  U: 4.8% (26) | Y: 64% (27)  N: 33% (14)  U: <5 | Y: 53.8% (34)  N: 42.0% (27)  U: <5 | Y: 50.8% (101)  N: 44.2% (88)  U: 5.0% (10) | Y: 35.4% (40)  N: 62.8% (71)  U: <5 | Y: 42.4% (48)  N: 52.2% (59)  U: 5.4% (6) |
| Interrupted family caregiving | Y: 47.5% (252)  N: 27% (143)  U: 25.5% (135) | Y: 52.4% (22)  N: 23.8% (10)  U: 23.8% (10) | Y: 42.8% (27)  N: 28.6% (18)  U: 28.6% (18) | Y: 49.2% (98)  N: 27.1% (54)  U: 23.6% (47) | Y: 40.7% (46)  N: 32.7% (37)  U:26.5% (30) | Y: 52.2% (59)  N: 21.2% (24)  U: 26.5% (30) |
| Disrupted home care | Y: 40.8% (218)  N: 30.1% (161)  U: 29.1% (156) | Y: 43.0% (18)  N: 30.9% (13)  U: 26.1% (11) | Y: 34.9% (22)  N: 34.9% (22)  U: 30.2% (19) | Y: 48.2% (96)  N:26.6% (53)  U:25.1% (50) | Y: 32.7% (37)  N: 38.0% (43)  U: 29.8% (33) | Y: 38.9% (44)  N:25.7% (29)  U: 35.4% (40) |
| Disrupted supplies | Y: 11.9% (64)  N: 37.1% (199)  U: 51.0% (274) | Y: 14.3% (6)  N: 40.5% (17)  U: 45.2% (19) | Y: 13.7% (8)  N: 38.1% (24)  U: 49.2% (31) | Y: 12.1% (24)  N:48.7% (97)  U:39.2% (78) | Y:8% (9)  N: 64.6% (73)  U:27.4% (31) | Y: 12% (14)  N: 52.2% (59)  U: 35.3% (40) |
| **Education System** | | | | | | |
| Excluded from in-person learning | Y: 14.4% (78) N: 43% (232)  U: 41.7% (225) | Y: 2.4% (1)  N: 73.8% (31)  U: 23.8% (10) | Y: 14.2% (9)  N: 38.1% (24)  U: 47.6% (30) | Y: 28 (14.1%)  N: 78 (39.2%)  U: 93 (46.7%) | Y: 11.5% (13)  N: 51.3% (58)  U: 37.1% (42) | Y: 19.4% (22)  N: 32.7% (37)  U: 47.8% (54) |
| Receipt of healthcare via education system | Y: 66.1% (357)  N: 11.5% (62)  U: 22.4% (121) | Y:76.2% (32)  N: 7.1% (3)  U: 16.6% (7) | Y: 58.7% (37)  N:14.3% (9)  U: 27% (17) | Y: 68.8% (137)  N:8% (16)  U:23.1% (46) | Y:62.0% (70)  N: 18.6% (21)  U:19.5% (22) | Y: 64% (73)  N: 22.1% (25)  U: 21.2% (24) |
| Transfer of resources | Y: 8.3% (45)  N: 31.3% (169)  U: 60.4% (326) | Y: 0% (0)  N: 38.1% (16)  U: 61.9% (26) | Y: 14.2% (9)  N: 19.0% (12)  U: 66.7% (42) | Y:9.5% (19)  N: 29.6% (59)  U: 60.8% (121) | Y: 8.8% (10)  N: 30.1% (34)  U: 61.2% (69) | Y: 8% (9)  N: 33.6% (38)  U: 58.4% (66) |
| Excluded because of PHA advice | Y: 8.3% (45)  N: 31.7% (171)  U: 60.0% (324) | Y: <5  N: 57.1% (24)  U:35.7% (15) | Y: <5  N: 34.9% (22)  U: 61.9% (39) | Y: 7.5% (15)  N:31.2% (62)  U: 61.3% (122) | Y: 6.2% (7)  N: 46.9% (53)  U: 46.8% (53) | Y: 7% (8)  N: 38.1% (43)  U:54.9% (62) |

*9 respondents did not provide province; we excluded NWT and Yukon because of small numbers

**Newfoundland and Labrador, Brunswick, Nova Scotia and Prince Edward Island areas Atlantic Canada, reflecting their unified approach to the COVID-19 pandemic.

*** Alberta, Manitoba and Saskatchewan were combined as The Prairies, reflecting the similarity of their pandemic experience.

Y: yes; N: no; U: unknown
