## Supplemental file 1 for "The impact of the COVID-19 pandemic on children with medical complexity"

CPSP
CANADIAN PAEDIATRIC SURVEILLANCE PROGRAM

### Survey

#### Impacts of the COVID-19 pandemic on children with medical complexity

**Your contribution is greatly appreciated.**

1. Do you provide care for children with medical complexity (CMC)* as part of your clinical practice?

   ** CMC have chronic conditions with high family-identified needs (e.g., need for substantial care in the home) associated with **fragility** (e.g., frequent hospitalizations), **technology-dependence** (e.g., oxygen, enteral feeding tube, tracheostomy), and the **need for multiple care providers** (e.g., specialists, community providers, and home care services). An example would be a child with a developmental disability, seizures and dystonia, requiring G-tube feeds, with two previous admissions to hospital in the past year. CMC include those not yet discharged from hospital (e.g., premature infants in the neonatal intensive care unit), but exclude children who have high family-identified needs without associated medical complexity (e.g., children with autism).*

   ❍ Yes ❍ No ***If no, thank you for completing the survey.***

2. Which of the following best describes your practice?
   ❍ General paediatrician ❍ Paediatric subspecialist, specify:__________________
   ❍ Other, specify: _____________________
3. What is your practice setting? (Select all that apply)
   a) ❑ Urban ❑ Suburban ❑ Rural/Remote
   b) ❑ Academic ❑ Non-academic
   c) ❑ Inpatient hospital ❑ Emergency/Urgent care centre ❑ Outpatient clinic ❑ Private office/Community setting
4. Provide the first three digits of the postal code of the practice where you spend the majority of your time: ___ ___ ___
5. How many years have you been in independent practice? _____ years
6. Approximately what proportion of your clinical time is dedicated to the care of CMC? _____ %

##### Healthcare delivery

7. Have you encountered CMC who experienced an adverse health outcome (e.g., preventable hospitalization) due to a COVID-19 pandemic-related disruption in healthcare delivery (e.g., interrupted primary care access)? ❍ Yes ❍ No
   a) *If yes*, how many?_______
   b) *If yes*, and you have encountered one patient only, please indicate all related outcomes with "1".
      *If yes*, and you have encountered multiple patients, please rank all related outcomes in order of frequency, starting with "1" = most common.
      For outcomes not encountered, please mark with "0".
      __ Hospital admission __ Extended hospital admission __ Unplanned surgery/Intervention __ ICU admission __Death
      __Loss of physical or developmental gains __ Other, specify: ________________________________
   c) Any additional comments? ________________________________________
8. Have any COVID-19-related changes in healthcare delivery benefitted CMC in your care? ❍ Yes ❍ No
   a) *If yes*, list beneficial changes: ________________________________

##### Family caregiving

9. Thinking about the families with CMC with whom you have interacted since the outset of the pandemic, have you encountered families with CMC whose **family caregiving** ability was significantly impacted by the COVID-19 pandemic (e.g., parental illness, loss of homecare)? ❍ Yes ❍ No ❍ Unknown
   ***If no or unknown,* proceed to question 10.**
   a) *If yes*, what percentage of families with CMC in your practice experienced significant challenges associated with family caregiving due to the COVID-19 pandemic? ❍ <20% ❍ 20–40% ❍ 41–60% ❍ 61–80% ❍ >80%
   b) What were the most frequent challenges faced by family caregivers? Rank the following in order from most frequent to least frequent, with "1" being most frequent. For challenges not seen, mark with a "0".
      __ Need for family caregiver self-isolation due to COVID-19 exposure or illness __ Family caregiver mental illness/burnout
      __ School/respite services closure (resulting in increased total number of weekly hours at home) __ Interrupted rehabilitation __ Loss of homecare __ Increased financial stress __ Other, specify: ________________________
   c) Any additional comments? ________________________________________

(…/2)

#### Homecare

10. Thinking about the families with CMC with whom you have interacted since the outset of the pandemic, have you encountered families with CMC who experienced disrupted **homecare** during the pandemic? ❍ Yes ❍ No ❍ Unknown

   ***If no or unknown,* proceed to question 11.**

   a) *If yes,* what percentage of families with CMC in your practice experienced disruptions in homecare delivery during the pandemic? ❍ <20% ❍ 20–40% ❍ 41–60% ❍ 61–80% ❍ >80%
   b) What were the most frequent homecare disruptions? Rank the following in order from most frequent to least frequent, with "1" being most frequent. For disruptions not seen, mark with a "0".
      __ Need for family self-isolation due to COVID-19 exposure/symptoms or infection __Decreased homecare availability
      __ Homecare worker illness or need for self-isolation __ Family choice to discontinue or limit homecare
      __Other, specify: ____________________________________________________________
   c) Any additional comments? _______________________________________________________

#### Medication and equipment supply

11. Did any CMC in your practice encounter difficulties accessing important supplies during the pandemic?
   ❍ Yes ❍ No ❍ Unknown

   ***If no or unknown*, proceed to question 12.**

   a) *If yes,* which important supplies were difficult to access? (Select all that apply)
      ❑ Medication ❑ Medical equipment ❑ Personal protective equipment ❑ Hand sanitizer ❑ Routine vaccinations
      ❑ Other, specify:__________________
   b) Specify if difficulties were due to (select all that apply):
      ❑ Medication or supply shortage; specify: ________________ ❑ Delays in prescribing; describe: ____________________
      ❑ Delays in dispensing; describe: _______________________ ❑ Other, specify: ___________________________________
   c) What was the most severe outcome associated with medication and equipment access difficulties? (Select one)
      ❍ Minor inconvenience (e.g., short delay, no change in care)
      ❍ Moderate issue (e.g., alternative medication or equipment sourced, no change in clinical status)
      ❍ Significant event (e.g., negative clinical outcome) ❍ None
   d) *If a significant event* was experienced by one of your patients, provide additional details: ______________________________
   e) *If a significant event* was experienced by one of your patients, did the negative clinical outcome result in (select all that apply): ❑ Hospital admission ❑ Unplanned surgery/intervention ❑ ICU admission
      ❑ Death ❑ Other, specify:_____________________________________
   f) Any additional comments? ____________________________________________________________________________________

#### Education, including virtual learning, school supports, in-school delivery of nursing and therapies

12. What percentage of school-aged CMC in your practice attended school in-person in September 2020?
   ❍ <20% ❍ 20–40% ❍ 41–60% ❍ 61–80% ❍ >80% ❍ Unknown
   a) What percentage of school-aged CMC in your practice attended school in-person in previous years (non-COVID-19 years)?
      ❍ <20% ❍ 20–40% ❍ 41–60% ❍ 61–80% ❍ >80% ❍ Unknown
13. Were CMC excluded from in-person learning when children without medical complexity were given the option to attend class?
   ❍ Yes ❍ No ❍ Unknown
   Any additional comments? _____________________________________________________________________________________
14. Do CMC in your practice typically receive any healthcare services via the education system (e.g., access to rehabilitation services, school nursing)? ❍ Yes ❍ No ❍ Unknown
   *If yes*, were such services transferred to home and/or community during periods of virtual learning? ❍ Yes ❍ No ❍ Unknown
15. Did public health advice preclude any children from attending school (e.g., children whose care involves aerosol generating medical procedures)? ❍ Yes ❍ No ❍ Unknown
   *If yes,* describe: _______________________________________________________________________________________________
16. Have you encountered issues relating to school closure that are particularly relevant to CMC (e.g., transportation)? ❍ Yes ❍ No
   *If yes*, describe: _______________________________________________________________________________________________
17. Any additional comments? _____________________________________________________________________________________

**Principal investigators:** Catherine Diskin, Peter J. Gill **Co-investigators:** Francine Buchanan, Eyal Cohen, Tammie Dewan, Tessa Diaczun, Michelle Gordon, Esther Lee, Nathalie Major, Charlotte Moore Hepburn, Julia Orkin, Hema Patel

**Please return this survey with your monthly reporting form. Thank you for your participation.**

02/2021
